# Characteristics, Interventions, and Deprivation Patterns in Suspected Opioid-Associated Out-of-Hospital Cardiac Arrests Attended by North East Ambulance Service NHS Foundation Trust: A 36-Month Service Evaluation

**DOI:** 10.64898/2026.10.20.26364948

**Authors:** Benjamin Kirk, Graham McClelland, Jason Scott

## Abstract

**Background:** Drug-poisoning deaths in England and Wales have more than doubled since 2012, with the North East of England persistently reporting the highest mortality rates. Opioid-associated out-of-hospital cardiac arrest (OA-OHCA) presents distinct clinical and operational challenges for ambulance services, particularly amid the emergence of potent synthetic opioids and uncertainty over the role of naloxone during cardiac arrest. This service evaluation sought to characterise the incidence, clinical features, and deprivation patterns of suspected OA-OHCA attended by North East Ambulance Service NHS Foundation Trust (NEAS), and to describe their geographic distribution.

**Methods:** A retrospective service evaluation was conducted using structured and reported fields from the NEAS electronic patient care record (ePCR) system between 1 January 2022 and 31 December 2024. Inclusion required a documented cardiac resuscitation together with either a reported aetiology of drug overdose, or documented naloxone administration. Data were analysed descriptively.

**Results:** 351 cases met the inclusion criteria. The mean age was 42.9 years; 69.2% were male. Cases represented a rising proportion of OHCA where resuscitation was attempted by NEAS across the period (4.7% in 2022 to 5.9% in 2024). Naloxone was administered in 87.2% of incidents, adrenaline in 86.9%, and amiodarone in 9.4%. Advanced airway management was recorded in 88.3% and intraosseous access in 75.5%; defibrillation was performed in 16.5%. Cases were concentrated in the most deprived communities, with Index of Multiple Deprivation (IMD) deciles 1–3 comprising 71.8% of the cohort.

**Conclusion:** Suspected drug-related cardiac arrests in the NEAS region predominantly affected working-age adults in deprived communities, and were managed with consistent airway, ventilation, vascular access, and naloxone administration alongside infrequent defibrillation, consistent with a predominantly asphyxial arrest model. This evaluation provides a descriptive baseline that could inform service planning and provides the empirical foundation for a subsequent qualitative study of paramedic clinical decision-making in OA-OHCA.

## Background

Drug-related mortality in England and Wales has continued on a long-term upward trajectory, with deaths related to drug poisoning reaching their highest recorded level in 2024, where 5,565 drug-poisoning deaths were registered, corresponding to 93.9 deaths per million people (Office for National Statistics, 2025). Opioids remain central to this pattern: 47.1% of all drug-poisoning deaths involved an opiate or opioid, and deaths involving synthetic opioids nearly quadrupled in a single year, rising from 52 to 195 between 2023 and 2024 (Office for National Statistics, 2025).

The North East remains the English region with the highest rate of drug-poisoning mortality, at 167.6 deaths per million - over two and a half times the rate recorded in the southern regions of England (Office for National Statistics, 2025). This persistent North-South divide in health outcomes has been documented for more than a decade (Bambra, Barr and Milne, 2014) and is consistent with the broader pattern of higher all-cause mortality and lower life expectancy observed across Northern regions (Kontopantelis *et al*., 2018).

Drug-related mortality follows a steep socio-economic gradient, with rates over five times higher in the most deprived areas than the least deprived (Office for National Statistics, 2025). These patterns are rooted in structural disadvantage: unemployment, housing insecurity, intergenerational poverty, and limited access to treatment services (Black, 2021). The increasing circulation of high-potency synthetic opioids substantially compounds this picture. Nitazenes, a class of highly potent synthetic opioids, and illicit fentanyl analogues are typically many times more potent than heroin; they are poorly detectable without toxicological analysis (Office for Health Improvement and Disparities, 2024). For ambulance services operating in high-prevalence regions, this represents a meaningful and evolving shift in the clinical profile of drug-related emergencies, and one that is inherently difficult to identify at the point of care. Diagnostic uncertainty is further compounded by polypharmacy, which is the norm rather than the exception in drug-related death: over half of drug-poisoning deaths nationally involve more than one substance, and it is frequently impossible to determine which agent was primarily responsible (Office for National Statistics, 2025).

This same pattern is evident in the North East of England, with an audit of drug-overdose cardiac arrests finding that 73% of cases had more than one drug recorded as suspected prior to arrest (Roebuck, 2022), and because opioids are commonly combined with benzodiazepines, pregabalin, gabapentin, and stimulants such as cocaine, an arrest attributed to drugs may reflect the combined or synergistic effect of several agents, only some of which are opioids.

Opioid-associated out-of-hospital cardiac arrest (OA-OHCA) is one of the most clinically distinct and potentially preventable forms of cardiac arrest encountered in the pre-hospital environment. Its primary mechanism is opioid-induced respiratory depression: activation of µ-opioid receptors in brainstem respiratory centres, particularly the pre-Bötzinger complex, progressively reduces respiratory rate, tidal volume, and chemosensitivity to hypercapnia and hypoxia (Eckert and Yaggi, 2022). The result is hypoventilation progressing to apnoea, with severe, sustained hypoxaemia driving subsequent cardiac arrest (Dezfulian *et al*., 2021).

While respiratory failure is the dominant mechanism, opioids can precipitate cardiac arrest through other pathways. Bacteraemia and systemic sepsis are recognised complications in people who inject drugs; methadone is associated with QTc prolongation and the risk of torsades de pointes; and direct pulmonary toxicity may contribute in some cases (Dezfulian *et al*., 2021). These secondary mechanisms are clinically relevant in patients with established dependency and are important considerations when managing what may superficially appear to be an OA-OHCA.

Clinical guidance for OA-OHCA reflects continuing uncertainty over naloxone’s role once cardiac arrest is established. The 2025 ILCOR Consensus on Science with Tretment Recommendations found insufficient evidence to recommend opioid-specific therapies, including naloxone, beyond standard resuscitation care, citing very low-certainty evidence from observational studies (Drennan *et al*., 2025; Grunau *et al*., 2025). This differs from respiratory and peri-arrest states, where naloxone administration remains supported as a good practice measure to reverse opioid-induced respiratory depression before cardiac arrest becomes established (Drennan *et al*., 2025).

United Kingdom guidance for ambulance services adopts a more permissive position. Historically, the Joint Royal Colleges Ambulance Liaison Committee (JRCALC) advised that naloxone may be administered where opioid involvement is suspected, reflecting the realities of pre-hospital uncertainty (JRCALC, 2020). Current JRCALC guidance frames cardiac arrest following opioid overdose as usually secondary to respiratory or hypoxic arrest, advises that naloxone may be considered where opioid toxicity is strongly suspected and should not delay other interventions, and notes that these patients may be managed on scene unless there is a reason to transport (JRCALC, 2026)

Within the North East Ambulance Service NHS Foundation Trust (NEAS), an agreed consensus of care for suspected overdose cardiac arrest is in place, and specifies administration of three doses of naloxone where opioids are suspected or confirmed (North East Ambulance Service NHS Foundation Trust, 2020). For paramedics working in high-prevalence areas, the divergence between international consensus, national guidance, and local protocol is not a theoretical debate: the questions of whether to administer naloxone, how much, and at what point in the resuscitation sequence must be resolved under significant time pressure and without toxicological confirmation.

Against this backdrop, this evaluation represents the first structured analysis of OA-OHCA within the NEAS region. Its aim was to characterise the incidence, clinical features and interventions delivered, as well as the geographic and deprivation patterns of suspected drug OHCA attended by NEAS over a 36-month period.

## Methods

### Design and setting

North East Ambulance Service NHS Foundation Trust (NEAS) is the statutory provider of emergency medical services for the North East of England, serving a geographically and socio-economically diverse region encompassing Newcastle upon Tyne, Gateshead, North and South Tyneside, Sunderland, Northumberland, County Durham, and the Tees Valley. The region includes dense urban centres, post-industrial towns, coastal communities, and remote rural localities. NEAS responds to approximately 440,000 emergency and urgent calls per year (North East Ambulance Service NHS Foundation Trust, 2026), and the region’s persistently high drug-poisoning mortality provides a critical context for this work. This retrospective service evaluation used routinely collected data from NEAS electronic patient care record (ePCR) system for all incidents occurring between 1 January 2022 and 31 December 2024. This 36-month window was selected to provide three complete calendar years of data, allowing examination of annual trends while remaining current at the time of analysis.

The project was registered and approved as a service evaluation by NEAS’ Clinical Effectiveness, Development, Audit and Research Group (CEDAR), and received approval from Northumbria University Ethics Committee (Project No. 8566). As a retrospective evaluation of existing service data, Health Research Authority approval was not required.

### Terminology

Throughout this paper, opioid-associated out-of-hospital cardiac arrest (OA-OHCA) denotes the clinical phenomenon under investigation. Where cases are described as ‘drug-related’ OHCA, this reflects a specific limitation of the data: opioid involvement could not be confirmed in every case. Cases were identified through documented naloxone administration or a reported aetiology of drug overdose, neither of which establishes with certainty that opioids were the causative agent. Naloxone is frequently administered empirically when drug involvement is suspected, irrespective of the substance, and a drug-overdose aetiology code does not distinguish opioids from stimulants, benzodiazepines, or poly-drug combinations. The variation in terminology is therefore deliberate: OA-OHCA is used where the discussion concerns the opioid-associated phenomenon specifically, and drug-related OHCA where the reference is to the broader cohort as defined by the inclusion criteria.

### Case identification and Selection

Cases were included where the following applied:

- Patients from any age group
- In cardiac arrest, with rationale this was secondary to drugs, defined as:

- Reported aetiology of “Drug overdose” AND/OR
- Documented naloxone administration.
- Received cardiac resuscitation (cardiopulmonary resuscitation (CPR), defibrillation, adrenaline, or amiodarone)

Advanced airway placement, ventilation, or vascular access alone did not qualify a case for inclusion, as these interventions may be undertaken during respiratory or peri-arrest presentations that do not constitute cardiac arrest. This cohort therefore comprises of patients in a suspected drug-related cardiac arrest in whom resuscitation was attempted.

Cases were excluded where no evidence of a resuscitation attempt was available - where no CPR, defibrillation, adrenaline, or amiodarone was recorded. In cases where the first and final observations showed absent perfusion and no recorded resuscitation interventions, the case was assumed to be Recognition of Life Extinct (ROLE) with no resuscitation attempted. Both were excluded, consistent with the exclusion of patients recognised as deceased on arrival. Duplicate records for a single incident were consolidated.

### Data sources and variables

All data were obtained from the NEAS ePCR system, supplied by NEAS Informatics. Variables comprised demographics (age, sex, postcode district, index of multiple deprivation, local authority), reported aetiology, interventions performed (manual and mechanical CPR, airway device, ventilation, intravenous and intraosseous access, defibrillation), medications administered, and incident outcome. The data were anonymised prior to release; no directly identifiable information was accessed. Geographic and deprivation indicators were derived from the incident-location postcode district already present in the extract. Because full postcodes were not released, small-area analysis was undertaken at postcode-district level.

### Analysis

Data cleaning, preparation, and analysis were conducted using LibreOffice Calc (The Document Foundation, 2025) and JASP (JASP Team, 2025). Analyses were descriptive. Continuous variables are summarised as mean (standard deviation) and range; categorical variables as counts and percentages. Case incidence was contextualised against the total number of resuscitation-attempted OHCAs attended by NEAS in each year, obtained from the Out-of-Hospital Cardiac Arrest Outcomes (OHCAO) registry reports (Brown, Booth and Perkins, 2023, 2024, 2025)

## Results

NEAS Informatics supplied 504 records representing 501 unique incidents of cardiac arrest patients which were administered naloxone or had a reported aetiology indicated drug overdose. Following consolidation of three duplicate records, and review against the inclusion and exclusion criteria, 351 unique incidents formed the final analytic cohort (Figure 1). Of the 150 incidents excluded, 140 had no documented cardiac resuscitation and were assumed to be Recognition of Life Extinct with no resuscitation attempted. The remaining ten had a viable perfusing output observation with no cardiac resuscitation delivered; the absence of arrest at the point of contact suggests these were more likely to represent respiratory arrests or peri-arrest presentations - a recognised feature of opioid toxicity - rather than true cardiac arrests, and they were excluded on this basis.

**Figure 1.**
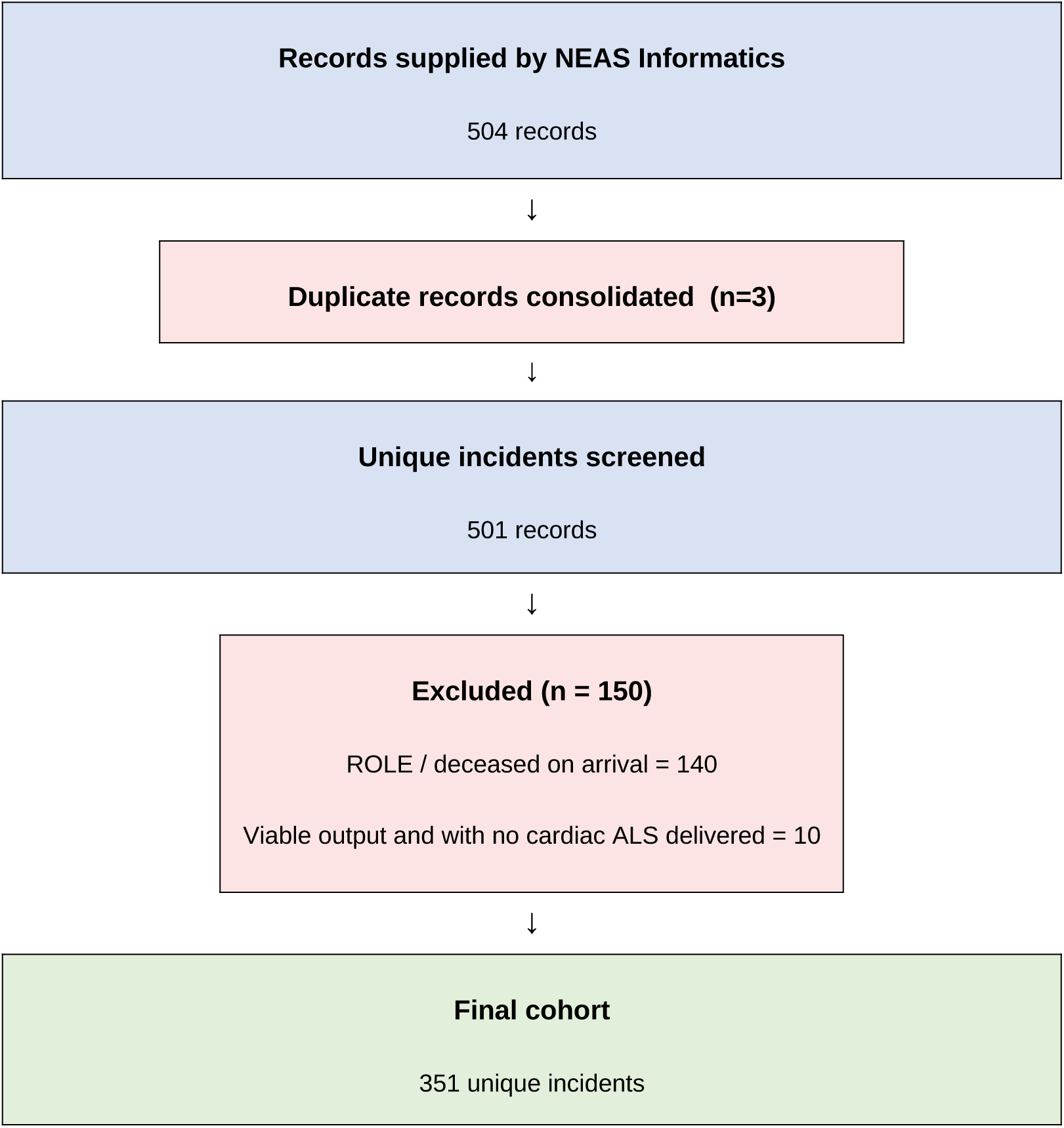
**Case screening and inclusion flow.**

### Demographic characteristics

The majority of patients were male (243/351, 69.2%); 107 (30.5%) were female and sex was not recorded in 1 (0.3%). The mean age was 42.9 years (standard deviation (sd): 13.6, range 14-91). The largest age group was 35-44 years (110 cases, 31.3%), followed by 45-54 years (91, 25.9%) and 25-34 years (69, 19.7%). Two cases (0.6%) involved patients under 18 years; these are retained in the cohort for completeness but, given their small number and distinct clinical context, are not analysed as a subgroup. The 25-54-year band accounted for 270 cases (76.9%). The mean age was similar between sexes, and the predominance of working-age adults is consistent with national drug-poisoning mortality data, in which the highest rates are observed among those aged 40-49 years (ONS, 2025). Ethnicity was recorded in only 65 cases (18.5%); it was documented as ‘unknown’ or not recorded in the remaining 286 (81.5%), with ‘White British’ the only substantive category among those recorded. This degree of incompleteness precluded any meaningful analysis of ethnicity, and no inferences regarding ethnic patterning of OA-OHCA can be drawn from these data.

**Table 1.** Demographic characteristics of the cohort.

| Characteristic | n = 351 (%) |
| --- | --- |
| **Sex** |  |
| Male | 243 (69.2) |
| Female | 107 (30.5) |
| Not recorded | 1 (0.3) |
| **Age band (years)** |  |
| <18 | 2 (0.6) |
| 18–24 | 23 (6.6) |
| 25–34 | 69 (19.7) |
| 35–44 | 110 (31.3) |
| 45–54 | 91 (25.9) |
| 55–64 | 29 (8.3) |
| 65+ | 24 (6.8) |

Mean age 42.9 years (SD 13.6, range 14-91), based on n = 348 valid ages (3 cases with missing data). Percentages of n = 351; may not sum to 100 due to rounding.

### Incidence and annual trend

OA-OHCA cases increased from 109 cases in 2022 to 114 in 2023 and 128 in 2024. Considered against the total resuscitation-attempted OHCA workload attended by NEAS, these represented a rising proportion of activity: 4.7% of 2,323 resuscitation-attempted OHCAs in 2022, 5.3% of 2,156 in 2023, and 5.9% of 2,180 in 2024 (Brown, Booth and Perkins, 2023, 2024, 2025) This proportional increase, rather than a simple rise in absolute counts, indicates that drug-related arrest is accounting for a growing share of the pre-hospital cardiac-arrest case-load.

### Geographic distribution and deprivation

Cases were distributed across all local authorities but clustered in a number of localities. The highest absolute numbers were recorded in County Durham (55, 15.7%), Northumberland (40, 11.4%), Sunderland (38, 10.8%), Newcastle upon Tyne (37, 10.5%), and Gateshead (35, 10.0%); together these five authorities contributed 205 cases (58.4%). When standardised to resident population, the ranking shifted: Redcar and Cleveland carried the highest three-year rate at 19.0 cases per 100,000, followed by Gateshead (17.8), Middlesbrough (17.4), South Tyneside (15.6), and Hartlepool (15.2). The prominence of Gateshead and several Teesside authorities in the population-standardised ranking reflects smaller resident populations bearing a disproportionate share of opioid-related harm rather than greater absolute numbers of cases.

Mapping at postcode-district level revealed marked spatial heterogeneity, with higher rates concentrated in urban and inner-city districts across the Tyne and Wear, and Teesside regions, with lower rates across suburban and rural districts. Cases were disproportionately concentrated in the most deprived areas: postcode districts in the most deprived decile (IMD decile 1) accounted for the largest share (140/351, 39.9%), and IMD deciles 1-3 together accounted for 252 cases (71.8%), while deciles 8-10 contributed only 27 (7.7%). OA-OHCA in this region is therefore not randomly distributed but occurs within identifiable geographic and socio-economic contexts.

**Table 3.** Distribution of OA-OHCA incidents by local authority (N = 351) Ranked by absolute case count. Population-standardised rates per 100,000 reorder this ranking (see text).

| Local authority | n (%) |
| --- | --- |
| County Durham | 55 (15.7) |
| Northumberland | 40 (11.4) |
| Sunderland | 38 (10.8) |
| Newcastle upon Tyne | 37 (10.5) |
| Gateshead | 35 (10.0) |
| Stockton-on-Tees | 28 (8.0) |
| Redcar and Cleveland | 26 (7.4) |
| Middlesbrough | 25 (7.1) |
| South Tyneside | 23 (6.6) |
| North Tyneside | 17 (4.8) |
| Hartlepool | 14 (4.0) |
| Darlington | 13 (3.7) |

### Geographic Distribution of Drug-Associated Out-of-Hospital Cardiac Arrests Attended by North East Ambulance Service NHS Foundation Trust, 1 January 2022 – 31 December 2024

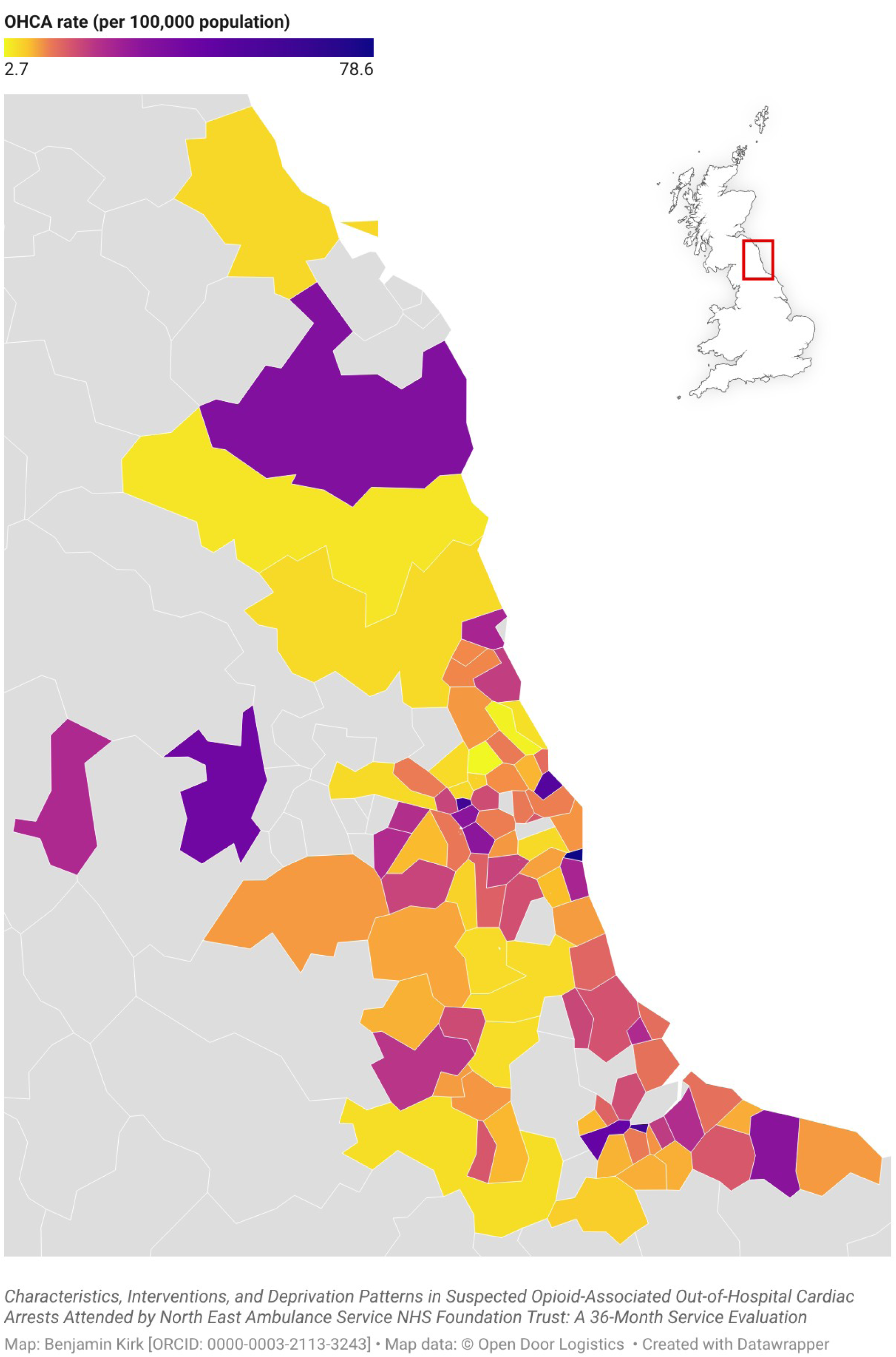

**Table 4.** Distribution of OA-OHCA incidents by Index of Multiple Deprivation (IMD) decile (N = 351)

| IMD decile (1 = most deprived) | n (%) |
| --- | --- |
| 1 | 140 (39.9) |
| 2 | 64 (18.2) |
| 3 | 48 (13.7) |
| 4 | 22 (6.3) |
| 5 | 24 (6.8) |
| 6 | 16 (4.6) |
| 7 | 9 (2.6) |
| 8 | 14 (4.0) |
| 9 | 8 (2.3) |
| 10 | 5 (1.4) |

### Clinical presentation

The recorded clinical presentation was dominated by cardiac arrest, which was documented as the presenting complaint in 250 cases (71.2%) and appeared in the clinical impression in the large majority. A minority presented with descriptors reflecting the peri-arrest or opioid-toxicity spectrum, including not breathing (12 cases, 3.4%), unresponsive (9 cases, 2.6%), and overdose (8 cases, 2.3%), with smaller numbers recorded as unconscious, seizure, or deceased. The impression field frequently combined *“cardiac arrest”* with an overdose descriptor, for example *“cardiac arrest with drug overdose”* (12 cases) or with *“intentional overdose”* (8 cases), reflecting the concurrent recognition of both the arrest and its suspected drug aetiology at scene. This distribution is consistent with a cohort identified at the point of established arrest, while the tail of respiratory and unresponsive presentations illustrates the continuum from opioid toxicity through respiratory compromise to cardiac arrest that characterises this patient group.

### Clinical interventions

A resuscitation attempt was documented in all included cases. Manual CPR was recorded in 286 cases (81.5%) and mechanical CPR in 79 (22.5%); these are not mutually exclusive, as mechanical devices may follow an initial period of manual compressions. Advanced airway management with an iGel supraglottic device was recorded in 310 cases (88.3%), and ventilation in 156 (44.4%). The discrepancy between airway placement and documented ventilation is notable: it is clinically implausible that ventilation was withheld in most cases where an advanced airway was inserted, and the gap most likely reflects inconsistent completion of the structured ventilation field rather than a true absence of ventilation. This represents an important data-quality limitation.

Intraosseous (IO) access was recorded in 265 cases (75.5%) and intravenous (IV) access in 231 (65.8%). Both routes were documented in 160 cases (45.6%), IO alone in 105 (29.9%), and IV alone in 71 (20.2%); no vascular access was documented in 15 (4.3%). Among pharmacological interventions, adrenaline was administered in 305 cases (86.9%) and amiodarone in 33 (9.4%). Defibrillation was performed in 58 cases (16.5%).

**Table 5.** Clinical interventions delivered during resuscitation (N = 351)

| Intervention | N (%) |
| --- | --- |
| Naloxone | 306 (87.2) |
| Adrenaline | 305 (86.9) |
| iGel (advanced airway) | 310 (88.3) |
| Intraosseous (IO) access | 265 (75.5) |
| Manual CPR | 286 (81.5) |
| Intravenous (IV) access | 231 (65.8) |
| Ventilation (structured field) | 156 (44.4) |
| Mechanical CPR | 79 (22.5) |
| Defibrillation | 58 (16.5) |
| Amiodarone | 33 (9.4) |

### Naloxone administration

Naloxone was recorded as administered in 306 of 351 incidents (87.2%). The two fields used to identify the cohort - recorded aetiology and naloxone administration - overlapped only partially. Naloxone and an overdose aetiology coincided in 126 incidents, whereas 180 (51.3%) received naloxone despite a recorded aetiology that was not overdose, most often "presumed cardiac" (114) and, in a substantial minority, asphyxial including hanging (55). In the remaining 45 incidents (12.8%), an overdose aetiology was recorded but no naloxone was documented. That naloxone was administered in over half of the cohort in arrests coded to a non-overdose aetiology is consistent with empirical treatment under diagnostic uncertainty, where opioid involvement cannot be excluded at scene rather than positively confirmed; it also means that surveillance relying on aetiology coding alone would substantially misclassify naloxone-treated arrests. Conversely, the 45 overdose-coded incidents without documented naloxone represent a quality-assurance signal warranting targeted case review; whether these reflect genuine non-administration, documentation omission, or suspicion of a non-naloxone-reversible substance cannot be determined from the available fields.

### Outcome and disposition

Return of spontaneous circulation (ROSC) was recorded in the structured impression field in only 3 of 351 incidents (0.9%), all among patients subsequently conveyed to hospital. In total, 166 incidents (47.3%) resulted in transport and the remaining 185 (52.7%) were managed on scene without conveyance, a disposition consistent with termination of resuscitation on scene in the large majority.

## Discussion

This service evaluation provides the first structured description of OA-OHCA epidemiology and management within a UK regional ambulance service. Over three years it characterises a consistent and proportionally increasing burden of suspected drug-related cardiac arrests, concentrated in working-age adults from the most deprived communities and managed with a pre- hospital intervention profile that reflects both the physiological demands of opioid-associated arrest and the practical realities of attending this population.

The demographic profile closely mirrors national drug-poisoning mortality data, supporting the cohort’s representativeness: the 69.2% male predominance aligns with the national figure of approximately two-thirds male drug-poisoning deaths, and the mean age of 42.9 years is consistent with national data identifying those aged 40–49 as highest-risk (Office for National Statistics, 2025). This convergence, together with the close correspondence to the CARES analysis of drug-overdose OHCA in the United States (Shekhar *et al*., 2024) suggests the structured fields, despite their limitations, capture a population that meaningfully reflects the wider epidemiology of opioid-related harm.

That cases represent a rising proportion of the resuscitation-attempted OHCA workload, not merely a rising count, is an important finding, because total OHCA workload was broadly stable across the period, the increase from 4.7% to 5.9% reflects a genuine shift in case-mix rather than a general rise in demand. This mirrors the trajectory reported from Arizona, where the proportion of overdose-related OHCA rose significantly over time and reached a comparable share of all treated OHCA (Smith *et al*., 2019) and it is consistent with the OHCAO registry’s own NEAS overdose-aetiology figures for the same years (Brown, Booth and Perkins, 2023, 2024, 2025).

Naloxone was documented in the large majority of arrests; however, its timing and sequence relative to other advanced life support interventions were not available. Given the continuing debate over naloxone in established cardiac arrest, it is important not to over-interpret its frequency as evidence of effectiveness: the data show how commonly it is used, but say nothing about whether it altered outcomes. What they do highlight is the need to understand decision sequencing - how clinicians balance airway management, ventilation, chest compressions, and other cardiac arrest medications against naloxone administration.

The intervention profile is consistent with a predominantly asphyxial arrest model. These findings extend a smaller local audit of drug-overdose cardiac arrest management undertaken within NEAS (Roebuck, 2022), which reviewed 139 cases attended between September 2020 and October 2021. That audit reported naloxone administration in 93% of cases where indicated, closely comparable to the 87.2% observed here across a larger cohort and later period, and it documented the same phenomenon of naloxone being administered even where opioids were not clearly suspected - the empirical, risk-averse pattern quantified in the present evaluation. The earlier audit also highlights the local protocol, in that three doses of naloxone should be administered while priority remains on airway management, ventilation, and early defibrillation, a pragmatic local adaptation of the JRCALC guidance. Critically, that audit’s attempt to test whether three-dose naloxone influenced return of spontaneous circulation (ROSC) was statistically non-significant but explicitly underpowered, and it concluded that evaluation with substantially more cases, and tighter control of confounding, would be required.

Beyond the aggregate intervention rates, the data reveals variation in how individual arrests were managed, consistent with clinical tailoring to the presentation rather than a single fixed algorithm. Naloxone and adrenaline were both documented in 274 cases (78.1%), while naloxone without adrenaline was recorded in 32 (9.1%) and adrenaline without naloxone in 31 (8.8%); this spread reflects the differing clinical judgements made at scene as to whether an arrest was principally opioid-driven, principally cardiac, or of uncertain cause. Resuscitation modality also varied, with mechanical chest compression devices deployed in 79 cases (22.5%) alongside predominantly manual compression elsewhere, and with vascular access achieved by differing combinations of intraosseous and intravenous routes. The variation in naloxone use relative to other interventions, in particular, underscores the value of the planned qualitative work in understanding how clinicians reason about these competing priorities under time pressure. The low defibrillation rate (16.5%) and low amiodarone use (9.4%) together indicate a predominance of non-shockable rhythms, aligning with the recognised pathophysiology of opioid-associated arrest (Dezfulian *et al*., 2021). However, in the absence of a structured rhythm field, this cannot be confirmed directly, nor can any subsequent rhythm changes be captured - for example, a shockable rhythm arising later in the resuscitation, or conversion following defibrillation or drug administration. The dataset therefore reflects only the interventions delivered, not the evolution of rhythm over the course of the arrest.

The high adrenaline rate (86.9%) is consistent with the inclusion criteria, which required evidence of cardiac life support, and confirms that the cohort comprised genuine cardiac arrests managed with advanced life support rather than peri-arrest presentations. The intraosseous access rate of 75.5% warrants particular comment: it is substantially higher than in general OHCA cohorts and reflects a familiar clinical reality - peripheral venous access is frequently impossible in people with a history of chronic injecting drug use, owing to progressive thrombosis, scarring, and obstruction of peripheral veins with repeated injection (Robertson, Broers and Harris, 2021). That both IO and IV access were recorded in 45.6% of cases is consistent with IO being used as rescue access following failed IV attempts. Recent RCT evidence indicates that, in OHCA more broadly, neither route shows consistent superiority in outcomes, supporting a pragmatic, flexibility-led approach to route selection rather than a fixed hierarchy (Saad *et al*., 2025).

Patterns by deprivation and geography reinforce national concerns about inequity in overdose burden. The concentration of cases in IMD deciles 1-3 (71.8%) is consistent with the steep socio-economic gradient in drug-poisoning mortality (Office for National Statistics, 2025). Gateshead’s position as the highest population-adjusted burden, despite fewer absolute cases than County Durham or Northumberland, is a reminder that raw counts can obscure the true burden carried by smaller, more deprived communities.

The poorly documeted ROSC rate almost certainly understates true ROSC: the ePCR impression field was not designed to capture patient outcome, and ROSC achieved during transport, recognised clinically but not documented, or attained after handover to hospital staff was completed, would all be missed. That only three events were captured across a cohort in which 166 patients were conveyed to hospital is itself difficult to reconcile with complete ascertainment. Because so few events were recorded, outcomes could not be analysed by deprivation, sex, or age; confirmed survival to discharge would require linkage to hospital records and was not available.

A recurring feature was the inconsistency between recorded aetiology and administered treatment, reflecting the reality that diagnostic certainty is frequently unavailable at scene. Where aetiology fields and intervention records are misaligned, administrative surveillance using aetiology coding alone will systematically undercount opioid-associated emergencies.

### Limitations

Several limitations should be acknowledged. The evaluation relied on structured and reported ePCR fields, so details of clinical presentation, overdose mechanism, and cardiac rhythm could not be confirmed. Intervention timing and sequencing were unavailable, and some interventions may be under-captured, as the ventilation data illustrate. Case ascertainment depended on naloxone administration or a recorded drug-overdose aetiology, so occult opioid-associated arrests attributed to other causes will have been missed, and opioid involvement could not be confirmed toxicologically in any case. The inclusion criteria required documented cardiac resuscitation (CPR, defibrillation, adrenaline, or amiodarone), consistent with the Utstein and CARES definition of an attempted resuscitation (Grasner *et al*., 2024; Cardiac Arrest Registry to Enhance Survival, 2025). This deliberately excluded cases managed with naloxone, airway, or vascular access without cardiac resuscitation, on the basis that these are consistent with respiratory or peri-arrest presentations rather than cardiac arrest. While this maximises the specificity of the cohort as a study of drug-related cardiac arrests in whom resuscitation was attempted, it is possible that a small number of genuine arrests with incomplete intervention documentation were thereby excluded, and the cohort does not capture opioid-associated arrests in whom resuscitation was not attempted. The screening and exclusion decisions, although documented in a transparent audit trail, were made by a single reviewer; independent second review would strengthen future work. Patient outcomes could not be robustly evaluated and would require linkage to hospital records. The findings should therefore be understood as an operational and epidemiological baseline rather than a clinical-outcomes analysis.

### Recommendations

The findings point towards several practical recommendations for NEAS and the wider system. First, the geographic and deprivation analyses make a clear case for targeted operational and harm-reduction response in the highest-burden localities, particularly Gateshead and the Teesside authorities; deployment, equipment, and skills should be weighted accordingly, and partnership with local-authority drug-treatment and take-home-naloxone services may reduce arrest incidence upstream. Second, the systematic inconsistency in aetiology coding is a data-quality concern with direct consequences for surveillance; a review of ePCR coding guidance for cardiac arrest would improve the reliability of administrative data. Third, given the rising proportion of the case-load and the growing penetration of synthetic opioids, front-line clinicians should have access to current guidance on OA-OHCA management, and any guidance review should specifically address synthetic-opioid recognition and naloxone dosing, since nitazenes and fentanyl analogues may require higher or repeated doses. Fourth, the high IO access rate supports routine availability of IO devices and maintenance of insertion competency, and prospective evaluation of IO-first access in this population is warranted. Fifth, prospective data collection incorporating intervention timing, cardiac rhythm, and linkage to hospital outcomes would substantially strengthen future evaluation.

### Conclusion

Across 2022–2024, North East Ambulance Service NHS Foundation Trust attended a substantial and proportionally increasing number of suspected drug-associated out-of-hospital cardiac arrests, predominantly in working-age male adults from the most deprived communities in one of England’s most socio-economically challenged regions. Care was characterised by consistent airway management, ventilation, vascular access, and naloxone administration alongside infrequent defibrillation, consistent with a predominantly asphyxial arrest. With drug-poisoning mortality still rising, synthetic opioids increasingly present, and cases already accounting for a growing share of the resuscitation caseload, there is no reasonable basis on which to expect this burden to diminish. This evaluation provides a robust descriptive baseline and the empirical foundation for a subsequent qualitative study of how experienced paramedics reason about, and manage, these complex and time-critical presentations.

## Data Availability

All aggregate data produced in the present study are contained in the manuscript. The underlying patient-level dataset is held by North East Ambulance Service NHS Foundation Trust and cannot be shared publicly; anonymised data may be made available on reasonable request, subject to NEAS Information Governance approval and an appropriate data sharing agreement.

https://www.neas.nhs.uk/contact-us/information-requests

## Acknowledgements

[author], NIHR Pre-doctoral Clinical & Practitioner Academic Fellow, is funded by the UK Department of Health and Social Care [grant ref]. The views expressed in this document are those of the author(s) and not necessarily those of the NIHR, Northumbria University, NHS, or the UK Department of Health and Social Care.

## 1. Declaration

The authors declare that they have no known competing financial interests or personal relationships that could have appeared to influence the work reported in this manuscript. No author has received payment, honoraria, consultancy fees, travel support, equipment, materials, drugs, writing assistance, gifts or other services from any commercial entity in connection with this work. No author holds patents, shares, or paid or unpaid positions in any organisation with a financial interest in naloxone, resuscitation equipment or the wider management of opioid-associated cardiac arrest.

## 2. Funding

This work was undertaken by BK during an NIHR Pre-doctoral Clinical and Practitioner Academic Fellowship, awarded through the NIHR Integrated Clinical and Practitioner Academic (ICA) Programme (Award reference: NIHR304863). The funder had no role in the design of the evaluation, the collection, analysis or interpretation of the data, the writing of the manuscript, or the decision to submit it for publication. The views expressed are those of the authors and not necessarily those of the NIHR or the Department of Health and Social Care.

## 3. Employment and institutional relationships

The following relationships are disclosed in the interests of full transparency. In the authors’ assessment none constitutes a competing interest capable of influencing the design, conduct, analysis or reporting of this evaluation.

**Benjamin Kirk** is a registered paramedic employed by North East Ambulance Service NHS Foundation Trust (NEAS), the ambulance service whose electronic patient care records form the dataset reported here, and is a Master of Research student at Northumbria University. He holds the NIHR fellowship described above.

**Graham McClelland** holds a joint appointment as Vice-Chancellor’s Fellow and Assistant Professor at Northumbria University, and as a research fellow at NEAS.

**Jason Scott** is Professor of Applied Health Research at Northumbria University and is a member of BK’s supervisory team. He declares no financial or non-financial interests beyond that academic appointment.

No author holds a managerial or clinical governance role at NEAS with responsibility for the care pathways described, and no author was involved in the clinical management of any case included in the analytic cohort in a capacity that could have influenced the data recorded.

## 4. Relationship with the host organisation

This work was conducted as a service evaluation of routinely collected NEAS data. NEAS is the data controller for the source records and granted access for the purposes of service evaluation. The evaluation received approval from Northumbria University (Project reference 8566); HRA and REC review were not required as the work constitutes service evaluation rather than research. NEAS placed no restriction on the reporting of findings, and the interpretation and conclusions presented are those of the authors alone.

## 5. Authorship

All named authors meet the ICMJE criteria for authorship, have approved the submitted version of the manuscript, and agree to be accountable for all aspects of the work. Individual contributions are set out in the accompanying CRediT author statement. There are no honorary or ghost authors, and no professional writing assistance was used in the preparation of this manuscript.

## CRediT Author Statement

**Benjamin Kirk:** Conceptualisation, Methodology, Formal analysis, Investigation, Data curation, Writing – original draft, review & editing, Visualisation, Project administration, Funding acquisition.

**Graham McClelland:** Conceptualisation, Methodology, Supervision, Project administration, Funding acquisition, Writing – review & editing.

**Jason Scott:** Conceptualisation, Methodology, Supervision, Funding acquisition, Writing – review & editing.

## Use of generative artificial intelligence

The authors confirm that generative artificial intelligence and AI-assisted technologies were not used to produce scientific or clinical content, to generate or alter data, or to draw conclusions. Any use was confined to language and formatting support under full author oversight, and all such output was reviewed and edited by the authors, who take full responsibility for the content of the publication.

## Notes

### Author Declarations

Northumbria University - Project No. 8566 Reference: Kirk 2025-8566-11499 Confirmation of Ethical Opinion The above application has now been approved via the Northumbria University Ethics Online System. The approval date is 19/05/2025.

